# Structured adult learning outside school or job associates with improved plasma markers of age

**DOI:** 10.64898/2026.01.11.26343888

**Authors:** William T. Hu, Michelle Migut, Hayley Horvath, Sarah Singer, Lisa Lanza Lopez, Ashima Nayyar, Mei-ling Li, Guibin Su, Stephanie Shiau, Stephanie Bergen, Margaret Koller, Benjamin Chapman, Paul Duberstein, Joel C. Cantor

## Abstract

**Background:** Peak educational attainment is linked to early life general cognitive abilities, mid-life socioeconomic status, and late-life dementia risks. Yet, it is difficult to study its mechanisms acting across decades of life given diverse paths towards peak attainment and lifelong learning opportunities.

**Methods:** We analyzed profiles of peak educational attainment, adult learning (formal, job-related, non-formal, informal), and their relationships to novel plasma proteomic markers of age in a regionally representative cohort of 580 adults.

**Results:** One in three participants obtained their highest degrees beyond first three decades of life, with differential associations with parental education and personality traits. Similar associations were observed for job-related, non-formal (structured learning outside of school or job), and self-directed informal learning. Only non-formal learning correlated with a protective profile of plasma proteins reflecting age-associated vascular dysfunction and arthritis.

**Conclusion:** Non-formal adult learning may improve plasma age markers and should be further investigated.

## Introduction

Education has been increasingly recognized as a critical determinant of brain health influencing cognitive function, neuroplasticity, and trajectories of aging.[1–7] Early life educational attainment and mid/late-life cognitive engagement are both associated with reduced risks for Alzheimer’s disease (AD) and related dementias[2, 8], but mechanistic evidence linking accomplishments and activities to brain aging or degeneration over decades is limited. Historically, researchers exploring the effects of cognitive reserve on aging trajectory have used years of formal education as a surrogate for the former. While effective, this neuro-centric concept of formal schooling overlooks factors underlying (e.g., paternal educational level,[9] childhood socioeconomic status [SES][10, 11] and adversities[12], possible genetic variants[13]) or complicating (e.g., sensorineural hearing loss,[14] delayed completion of formal schooling beyond the third decade of life[15]) educational levels. Educational attainment is also but one element of lifelong learning which encompasses early childhood knowledge/skill acquisition, adulthood exposures/experiences, and ongoing intellectual engagement. At the societal level, the Organization for Economic Cooperation and Development (OECD) broadly categorizes a population’s opportunities for lifelong learning into three subtypes: formal (structured learning associated with a degree/certificate program), non-formal (structured learning outside of formal learning, such as swimming lessons or non-credit adult education courses), job-related (which has undergone transition from non-formal to more formal), and informal (non-structured learning through everyday experience or self-directed knowledge acquisition without an instructor).[16, 17] Lifelong learning is thus not only an individual achievement, but also a population-level factor with impact on cognitive health outcomes and societal well-being.

Instead of an active force capable of shaping cognitive reserve, educational attainment can instead be a passive reflection for general cognitive abilities (GCA)[18]. Recent work in the Vietnam Era Twin Study of Aging, Danish Conscription Database, and Swedish Screening Across the Lifespan Study found educational attainment to have limited impact on later dementia risks after adjusting for GCA.[18–20] If educational attainment’s reduction in dementia risks is mostly explained by GCA, formal learning in mid- and late-life[21] may neither expand cognitive reserve nor reduce dementia risks. In keeping with this, higher educational attainment is linked to better childhood SES, which in turn associates with parental educational levels.[22–27] Mid- and late-life learning opportunities have similarly been associated with early[28] or later life SES,[29, 30] while studies on intergenerational socioeconomic mobility and cognition have resulted in mixed findings. [31–35] Complicating these analyses is the temporal trend of rising educational attainment. In the United States (U.S.), proportion of adults 25 years and older having received a bachelor’s degree rose from 21.3% in 1990 to 37.5% in 2020[36]. This near doubling is not likely explained by similar improvement in cognitive abilities,[37, 38] and Americans who complete higher education later in life differ in race/ethnicity and sex from those who complete the same degrees earlier[39]. Thus, lifetime peak educational attainment is influenced by social factors as well as GCA, and separately examining the effects of educational attainment during and lifelong learning after active periods of brain development[40] could better inform interventions and policies.

We hypothesized that peak educational attainment and lifelong learning patterns differed according to age, sex, race/ethnicity, parental education, and personality traits in a contemporary cohort. We tested this hypothesis by detailed assessment within the Lifelong Learning Sub-study (LLS, n=580) within the larger New Jersey Population Health Cohort (NJHealth; n=2,503 as of 6/1/2025, with median age 56 and interquartile [IQR] range of 38-69) Study. NJHealth has a highly educated participant base, with over 60% having at least a bachelor’s degree (vs. 27% of their mothers and 35% of their fathers) which outpaces the national trend[36]. Coupled with the racial/ethnic diversity in NJHealth (43.5% reporting as non-Hispanic White [NHW]), this regional cohort potentially represents a behavioral, social, and biomedical forecast into a more educated, majority-minority U.S. population. To further explore lifelong learning’s potential mechanistic effects on cognitive aging and dementia risks, we also examined their relationship to novel age-associated markers to identify activities most likely to exert a measurable biological outcome.

## Methods

### Ethics Approval

This study was approved by the WCG Institutional Review Boards (IRB). All work was planned and conducted in concordance with the WMA’s Declaration of Helsinki as revised in 2024.

### Consent to Participate

Signed and dated informed consents were obtained from all participants.

### Study Design and Recruitment

NJHealth is longitudinal and on-going with a focus on life course factors’ health implications. It combines probability-based sampling (59%) for representativeness throughout the US State of New Jersey (NJ) with respondent-driven sampling for difficult-to-reach immigrant populations.[41] In the parent study, NJ residents aged 14 and older were recruited using probabilistic and purposive methods to include members of multigenerational families, marginalized populations, and recent immigrant groups. Study procedures included interviews (medical co-morbidities, social and migration history, self-reported individual and societal stressors, associated stress buffers and amplifiers), health and well-being outcomes (cognitive assessment, activities of daily living, depression, anxiety, discrimination-related, suicide-related), salivary or peripheral blood DNA, and fasting (when convenient and appropriate for health reasons) plasma collection. All research procedures take place at participants’ homes or at Rutgers Health in New Brunswick. Participants receive $55 total compensation for completing all procedures, or prorated for activities completed. For LLS, a cross-sectional design was used to report the association between parent study’s baseline findings and additional data on formal, job-related, non-formal, and informal learning among a conveniently sampled subgroup (n=580, Table 1). Compared to the remaining NJHealth participants, LLS participants were more likely to have been recruited through probability sampling (85.0% vs. 51.6%), NHW race/ethnicity (70.3% vs. 35.4%), older age (median age 60 vs. 53), health insurance, preventive health services, and reported volunteering hours (Table 1).

**Table 1.** Demographic, medical, social, and behavioral characteristics of Lifelong Learning Sub-study (LLS) participants and the plasma marker validation cohort. .

|  | **LLS**<br>**(n=580)** | **Non-LLS**<br>**(n=1,923)** | **p** |
| --- | --- | --- | --- |
| Population-based sampling | **493 (85.0%)** | 993 (51.6%) | <0.001 |
| Female | 360 (62.1%) | 1237 (64.3%) | 0.321 |
| Age | **60 (42-69)** | 53 (35-68) | <0.001 |
| Educational level (yrs equivalent for degree) | **16 (16-18)** | 16 (14-17) | <0.001 |
| BMI (kg/m<sup>2</sup>) | **27.2 (24.2, 31.1)** | 25.7 (22.7, 29.3) | <0.001 |
| Marital Status |  |  | 0.045 |
| Single, never married | 129 (22.2%) | 528 (27.5%) |  |
| Married | 317 (54.7%) | 982 (51.1%) |  |
| Widowed, separated, divorced | 134 (23.1%) | 413 (21.4%) |  |
| Race/ethnicity |  |  | <0.001 |
| Hispanic American | 60 (10.3%) | **317 (16.5%)** |  |
| Black/African American | 37 (6.4%) | 160 (8.3%) |  |
| Asian American | 62 (10.7%) | **711 (37.0%)** |  |
| Other/Multiple | 13 (2.2%) | 54 (2.8%) |  |
| White | **408 (70.3%)** | 681 (35.4%) |  |
| **Parental educational level** |  |  |  |
| Father with a Bachelor's degree | 211 (36.4%) | 664 (34.5%) | 0.260 |
| Mother with a Bachelor's degree | 168 (28.9%) | 498 (25.9%) | 0.142 |
| **Medical History** |  |  |  |
| Cardiovascular |  |  |  |
| Hypertension | 236 (40.6%) | 718 (37.3%) | 0.145 |
| Hyperlipidemia | **324 (55.9%)** | 893 (46.3%) | <0.001 |
| Limb claudication | 76 (13.1%) | 266 (13.8%) | 0.654 |
| Diabetes | 71 (12.2%) | 228 (11.9%) | 0.802 |
| Smoking/Ex-smoker | **112 (19%)/57 (10%)** | 330 (17%)/138 (7%) | <0.001 |
| Myocardial infarction | 32 (5.5%) | 101 (5.2%) | 0.264 |
| Atrial fibrillation | 47 (8.1%) | 117 (6.1%) | 0.085 |
| Chronic kidney disease | 21 (3.6%) | 71 (3.7%) | 0.936 |
| Exposure to statin | **211 (36.4%)** | 517 (26.9%) | <0.001 |
| Respiratory |  |  |  |
| Asthma | **51 (8.8%)** | 94 (4.9%) | <0.001 |
| COPD | 23 (4.0%) | 75 (3.9%) | 0.943 |
| Sleep apnea | **87 (15.0%)** | 146 (7.6%) | <0.001 |
| Neurological |  |  |  |
| Migraine | 70 (12.1%) | 190 (9.9%) | 0.130 |
| Seizure | 13 (2.2%) | 46 (2.4%) | 0.834 |
| Stroke | 18 (3.1%) | 85 (4.4%) | 0.162 |
| Mild TBI | 65 (11.2%) | 199 (10.3%) | 0.555 |
| Dream enactment | 32 (5.5%) | 85 (4.4%) | 0.273 |
| Having had COVID-19 | **394 (67.9%)** | 1193 (62.0%) | 0.010 |
| Arthritis | **235 (40.5%)** | 521 (27.1%) | <0.001 |
| Insomnia | 160 (27.6%) | 486 (25.3%) | 0.264 |
| History of cancer | **106 (18.3%)** | 201 (10.5%) | <0.001 |
| PHQ-8 | 3 (1-6) | 3 (1-6) | 0.212 |
| GAD-7 | 3 (1-6) | 3 (0-6) | 0.512 |
| UCLA Loneliness Scale-3 | 4 (3-5) | 4 (3-5) | 0.631 |
| **Health insurance** | **559 (96.4%)** | 1,730 (90.0%) | <0.001 |
| ADI |  |  |  |
| National rank (percentile) | 27 (16-39) | 26 (15-41) | 0.800 |
| State rank (decile) | 5 (3-7) | 5 (3-8) | 0.501 |
| **Preventive Care** |  |  |  |
| Flu vaccine | **404 (69.7%)** | 1,173 (61.0%) | <0.001 |
| Colonoscopy | **374 (64.5%)** | 1,013 (52.7%) | <0.001 |
| **Religious attendance** |  |  | <0.001 |
| Weekly or more | 160 (27.6%) | 698 (36.3%) |  |
| Monthly/A few times a year | 123 (21.2%) | 481 (25.0%) |  |
| Seldom or never | 297 (51.2%) | 744 (38.7%) |  |
| **Weekly volunteer hours** | **1 (0-2)** | 0 (0-2) | <0.001 |

### Questionnaires

As part of the broader NJHealth questionnaire, participants provided information on sex/gender; detailed race/ethnicity according to major groups in NJ including Hispanic (Mexican/Chicano, Puerto Rican, Cuban, Dominican, Other), non-Hispanic Asian (Chinese, Korean, Asian Indian, Japanese, Filipino, Vietnamese, Other), non-Hispanic Black/African American, non-Hispanic White, Multiple, and Other; address to calculate Area Deprivation Index(ADI)[42]; marital status; health insurance status; medical co-morbidities related to cardiovascular (hypertension, hyperlipidemia, diabetes, atrial fibrillation, myocardial infarction, limb claudication, chronic renal insufficiency or failure, exposure to statin therapy), respiratory (chronic obstructive pulmonary disease [COPD], sleep apnea, current asthma), neurological (stroke, migraine, seizure, mild traumatic brain injury [TBI], dream enactment) functions as well as history of cancer, insomnia, arthritis, COVID-19, and symptoms suspected to represent long COVID. Participants completed surveys to assess personality (Big Five),[43] depression (Patient Health Questionnaire-8, PHQ-8), anxiety (General Anxiety Disorder-7, GAD-7), loneliness (short UCLA scale of 3 questions designed for large surveys, ULS-3),[44] adverse childhood experiences (ACEs),[45] past trauma (Trauma History Questionnaire, THQ), and ADL (Katz Index of Independence of ADL). Participants also provided information on work status (employed for wages or self-employed, retired or unable to work, out of work, homemaker, student), attendance of religious services (never, seldom, a few times a year, once or twice a month, once a week, more than once a week), volunteering hours (average weekly time rounded to nearest integer, 0-5), subjective report of having a foreign accent (none, light, moderate, heavy), and participating in caregiving within the past 30 days.

Relevant to education, the original NJHealth surveys included questions on participants’ own and parental educational levels modeled after the Health and Retirement Study with greater details among those without than those with a bachelor’s degree, with no distinction between those with master’s and doctorate degrees[23, 46]. For more detailed assessment of educational level and lifelong learning, 580 participants (23% of NJHealth) completed the LLS. LLS assessed highest degree obtained, age of completion at educational milestones (6^th^ grade, high school diploma or passing the General Education Development Test [GED], first bachelor’s degree, first master’s degree, first doctorate, most recent degree), Likert scale for frequency (daily, weekly, a few times a year, 1-2 times a year, never) of formal learning (with an instructor, in a class format) as part of a degree/certificate program, job-related learning (historically considered non-formal but has become more formal[16]), non-formal learning (taking a class in a structured format, not as part of a degree/certificate program or as a part of job-related training), informal learning (unstructured knowledge/skill gathering or enrichment undertaken by the participant).

### Plasma collection and marker analysis

A custom assay of 100 inflammatory proteins and, when known and available, their antagonists as well as key markers for physiologic functions (cystatin C for glomerular filtration rate) were selected to undergo technical validation at two vendors chosen from a larger pool for willingness to disclose freeze-thawing information related to analysis (Supplementary Methods). The final panel contained 100 analytes measured by aptamer-based assays (SomaLogic, Boulder, CO) with well-characterized performance characteristics (intermediate precision; intra-individual biological variability; effects from freeze-thawing or delay in processing). All blood samples were always immediately processed to generate plasma aliquots (<30 minutes from arm to freezer), randomized, blinded, and shipped on dry ice without additional thawing (e.g., for sub-aliquoting).

We additionally compared 15 participants’ analyte level variability over three weeks after overnight fasting, and between collection without overnight fasting and average values from the three post-fasting collections. 11 analytes had greater CV between fasting and non-fasting conditions than among the three fasting draws, and only one’s difference was more than 10% (BPI, 3.1% vs. 2.5%; Table S3). Thus, fasting was encouraged but not required for NJHealth participants, for whom approximately 30 mL of blood was collected on the same day in K_2_-EDTA tubes, centrifuged at 2000 g and 4 °C for 5 min before aliquoting, labeling, and freezing at -80°C until analysis.

## Quantitation and Statistical Analysis

### Missing Data

In NJHealth, 33 of 2,503 participants (1.3%) provided addresses which could not be geocoded for ADI determination and were excluded from analyses involving ADI. Declined responses in ACEs were treated as positive responses to calculate total ACEs due to previous report of shared demographic variables and health indicators between non-responders and affirmative responders.[47] Ten participants in NJHealth (one in LLS) reported having volunteered but did not report the average volunteering hours per month. These missing values were replaced by the mean value from the overall group (4 hours per month). Eight LLS participants did not provide age for completing any of the educational milestones and were excluded from analyses involving these variables. Nineteen LLS participants did not provide age at which they completed 6^th^ grade but did provide age for later milestones.

### Analysis of baseline characteristics

All demographic and clinical data were analyzed using IBM SPSS 30 (Aramonk, NY). For baseline comparisons between those who provided or did not provide detailed life learning information, Student’s T-test or analysis of variance (ANOVA) were used for continuous variables, while Chi-squared tests were used for categorical variables. ULS-3 scores were analyzed as a dichotomous variable (6-9 as lonely). PHQ-8 and GAD-7 were analyzed after log-transformation due to positively-skewed distribution. National ADI also showed a positively-skewed distribution which normalized through square root transformation. Among BigFive personality scores, openness, agreeableness, and conscientiousness all showed negative skew while neuroticism showed a positive skew. BigFive traits were thus examined according to tertiles.

### Early life and lifelong learning activity analysis

To differentiate between early and mid/late-life attainment of highest educational degrees, age at obtaining each degree (high school/GED, associate, bachelor’s, master’s, doctorate) within LLS was used to derive two variables: highest educational attainment before age 30, and degree obtained after 30 (categorized as none, n=408; up to bachelor’s, n=37; graduate degree, n=127). Frequency of each lifelong learning activity among those 30 years or older was then entered into an ordinal regression model, with stepwise introduction of independent variables including age^2^ or age, sex, marital status, race/ethnicity, educational attainment before 30, degree obtained after 30, paternal highest degree, maternal highest degree, BigFive personality tertiles, medical co-morbidities, PHQ-8, GAD-7, being lonely, ACEs, and square root of national ADI (hereby referred as ADI in text but sqrt[ADI] in tables). Based on gender roles in the U.S., we also included the interaction terms between degree obtained after 30 and sex or marital status. Non-formal and informal learning models incorporating all five Likert responses met the parallel line assumptions, while degree/certificate program-related learning incorporating three Likert response (never, 1-2 times or a few times a year, weekly or daily) also met the assumption. Proportional odds for greater frequency were reported for each factor. Job-related learning failed the parallel line assumption, and was first analyzed in a multi-nominal logistic regression model which distinguished those with weekly or more job-related learning from the rest. The final job-related learning variable was thus analyzed using a binary logistic regression model, and odds ratios for greater frequency were presented for each factor.

### Plasma analyte measurement and analysis

Plasma analyte levels were processed through a standardized workflow to account for normality, inter-plate variability, outliers, and scaling (see Methods and Supplementary Methods). Following our own prior workflow as well as consensus recommendations[48, 49], dimension reduction was achieved via Principal Component Analysis (PCA). Briefly, each individual plasma analyte was first examined for distribution across the cohort using Kolmogorov-Smirnov Test, and analytes which did not have normal distribution were log10-transformed. After log10-transformation and standardization, outliers (≥5.0 or ≤-5.0) were removed and analytes were re-standardized before removed outliers were imputed in SPSS using five nearest neighbors. PCA was conducted using co-variance matrix and Varimax rotation, and the optimal number of principal components (PCs) was selected based on a combination of eigenvalue (> 0.70), elbow rule, and amount of variation explained (∼75%).

To identify plasma markers associated with age,[50] PC scores were analyzed using Pearson’s correlational analysis with age as well as age squared (age^2^), with Hochberg correction of the Bonferroni methods to account for multiple comparisons. Linear regression models were then used with these age-related PC scores as dependent variables, with age or age^2^, sex, race/ethnicity, medical co-morbidities, PHQ-8, and GAD-7, ACES, and ADI as independent variables to provide putative disease-related description of each age marker sets.

### Association between age-related PC scores and lifelong learning activities

Age-related PC scores’ associations with one’s own educational attainment before age 30, degree at 30 years of age or older, lifelong learning activities (any vs. none for each subtype), ADI, and parental educational attainment were examined by introduction into the demographically and medically adjusted regression models above. If more than one term was associated with a PC score, the model was examined for co-linearity. Lifelong learning activity associated with age-related PC scores were then examined for possible dose effect, including grouping of Likert scale responses according to similar regression coefficients and/or significance levels. Finally, lifelong learning activities’ associations with age-related PCs were examined for possible mediation effect by other measures of social engagement (frequency of religious attendance, average number of volunteer hours) or other needs for structured learning (having a foreign accent, being a caregiver)

### Availability of Data and Materials

Datasets used and/or analyzed during the current study are available from the corresponding authors on reasonable request according to data-sharing policies approved by Robert Wood Johnson Foundation and Rutgers University.

## Results

### Lifetime educational attainment extends into mid- and late-life

Within LLS (n=580), there was a much wider range of age at obtaining master’s or doctorate degrees (IQR of 11 and 14 years) than high school/GED or bachelor’s degrees (IQR of 2 years each, Fig 1A). Approximately one in three (31%) obtained their highest degree at 30 years of age or older, including over 40% of those with master’s (65/157) or doctorate (29/64) degrees (Fig 1B). Multi-nominal logistic regression analysis showed largely different factors promoting completion of up to a bachelor’s degree vs. a graduate degree at 30 years or older (Table S3). Specifically, intermediate agreeableness, reporting one ACE, and maternal education beyond high school/GED were all associated with increased odds of completing high school/GED, associate degree, or bachelor’s degree. Conversely, reporting one ACE reduced the likelihood of completing a graduate degree at 30 or older, along with intermediate or high neuroticism, and father having less education than a graduate degree. Thus, while often conceptualized as a singular early life process in life course aging studies or as a surrogate for GCA, lifetime educational attainment extends well into mid/late-life according to childhood adversity, parental education, and personality in this contemporary cohort.

**Figure 1.**
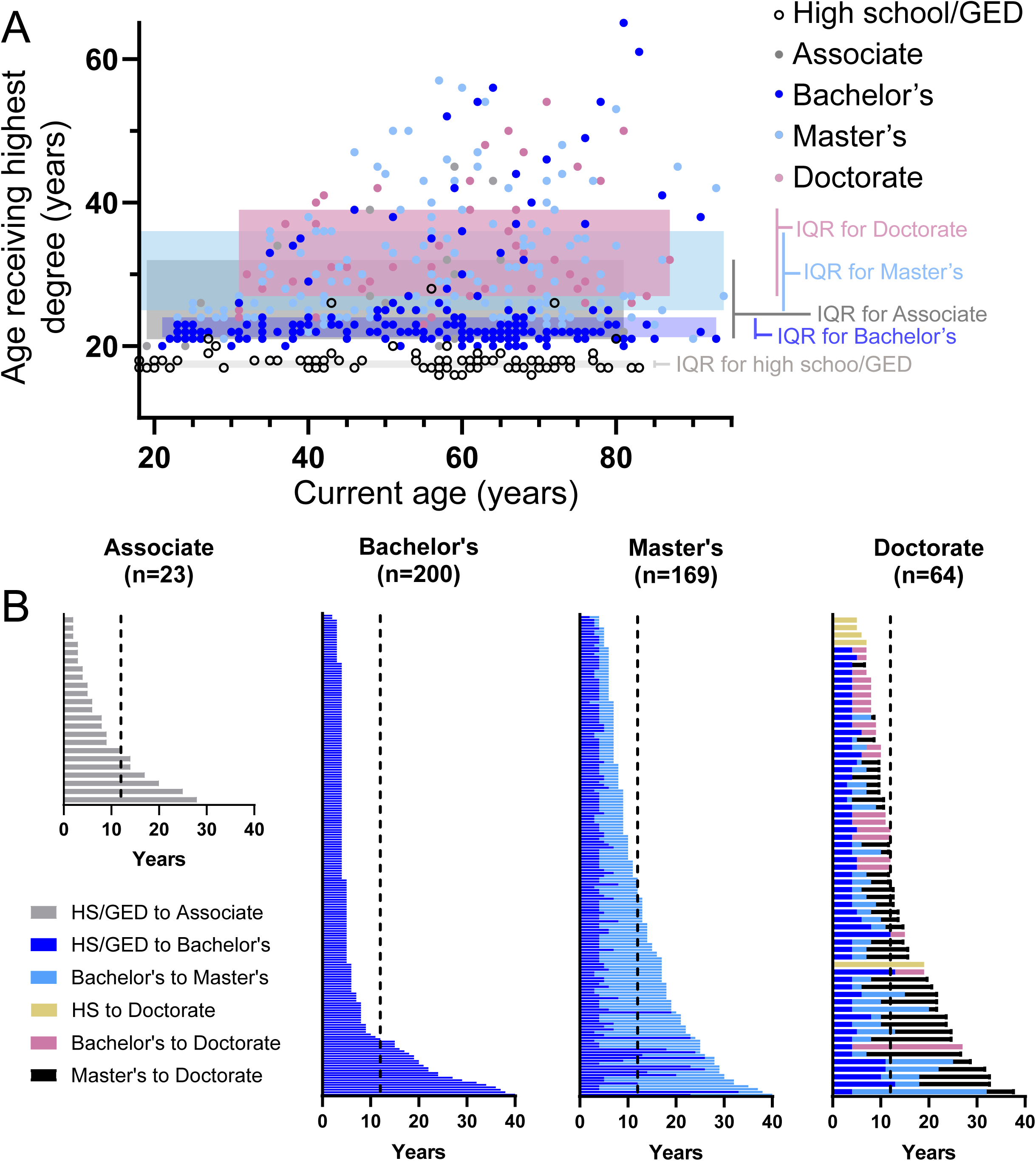
Variability in age at completion of highest degree among LLS participants. Whereas most participants completed high school/GED or bachelor’s degrees as their highest degrees before 25, those who obtained associate, master’s, or doctorate degrees did so over much wider age ranges (A; IQR=Interquartile range). When participants were examined according to highest degree attained (B), those with master’s degrees graduated college at a younger age than those with bachelor’s degrees (p=0.003; dashed lines represent age 30 based on median age of completing high school [HS] or GED).

### Factors associated with frequency of lifelong learning activities

In LLS, informal learning (self-directed knowledge/skill improvement) was the most commonly reported type of adult activity, with only 18% of respondents reporting no such activity within the past year. Formal learning as part of a degree or certificate program was most common in the youngest age group, while job-related learning was more common in the 30-49 age group (Fig 2A). 69 (12%) reported not having participated in any form of lifelong learning activity within the past year.

**Figure 2.**
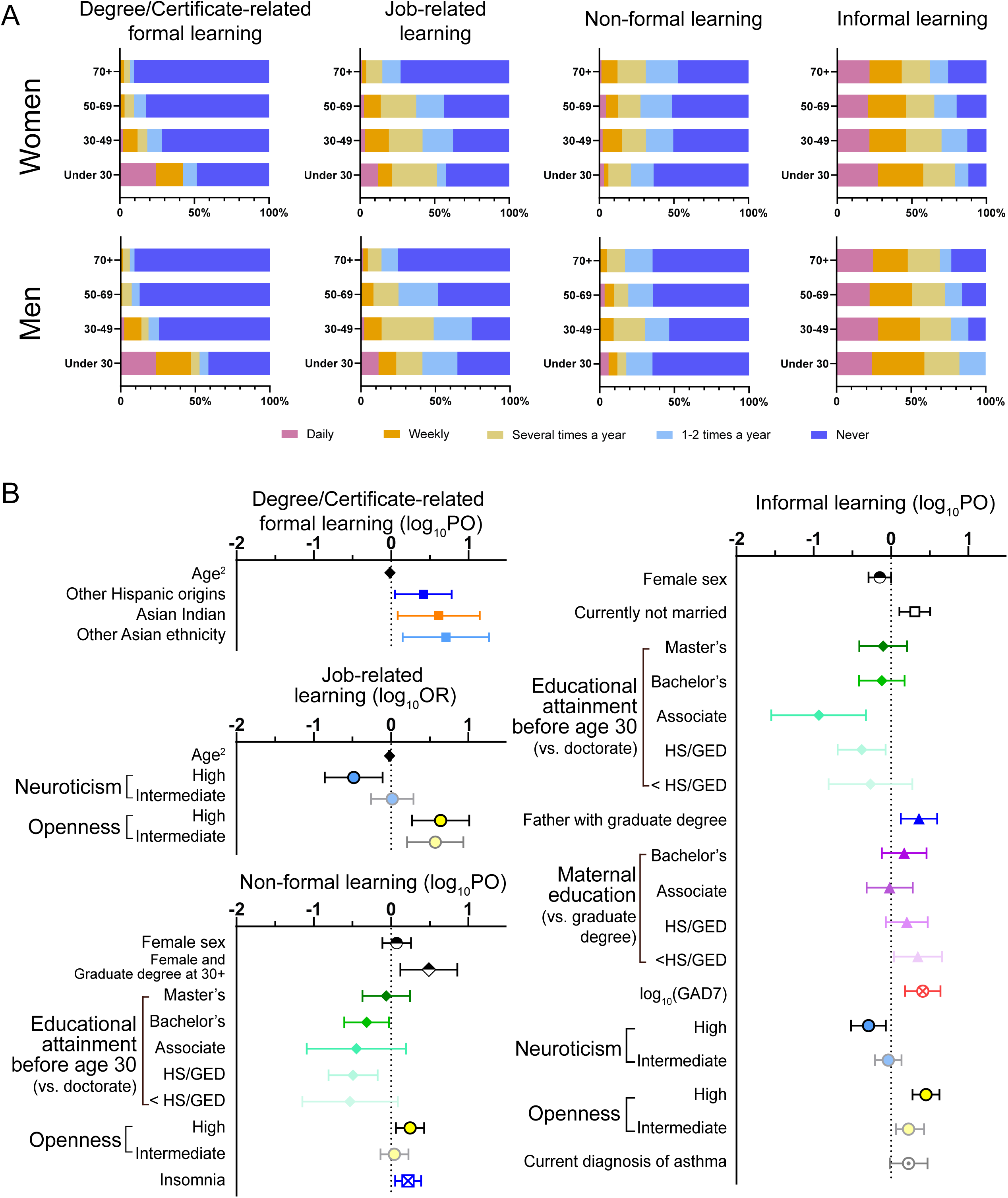
Lifelong learning activities of LLS participants. Participation in degree/certificate-related formal learning, job-related learning, non-formal learning, and informal learning differed according to age and sex (A). However, ordinal (proportion odds [P.O.] shown) or binary logistic regression (odds ratio [O.R.] shown) analyses among those 30 years or older showed one’s own educational attainment, parental educational attainment, and personality traits to consistently associate with multiple forms of lifelong learning (B). P.O. and O.R. are shown after log_10_-transformation for ease of interpretation.

To better identify factors which may contribute to greater frequency of each lifelong learning activity, we performed ordinal regression analysis for degree/certificate-related formal learning, non-formal learning, and informal learning, followed by logistic regression for job-related learning due to failed parallel line assumption (see Methods). Because formal education is generally completed during the second and third decades of life, we excluded people under 30 years of age (n=50, 8.6%) for subsequent analyses. Among the remaining LLS participants, degree/certificate-related formal learning was still associated with younger age as well as three ethnic backgrounds: Hispanic other than Mexican/Chicano, Puerto Rican, or Dominican; Asian Indian ethnicity; and Asian ethnicity other than Chinese, Korean, and Asian Indian (Fig 2B, Table S4). Younger age was also associated with increased odds of having job-related learning more than a few times a year (Table S5), but we found personality traits to more strongly influence multiple forms of lifelong learning. Specifically, greater openness[51] was associated with more frequent participation in job-related learning, non-formal learning, and informal learning, while less neuroticism was associated with increased participation in job-related learning and informal learning (Fig 2B, Tables S5-S7).

Several associations were shared by non-formal and informal learning, although not always in the same pattern (Fig 2B, Tables S6 & S7). Frequency for both types of learning increased with greater educational attainment before 30 years of age and having certain health challenges (insomnia for non-formal learning; asthma and anxiety for informal learning). On the other hand, women were more likely to undertake non-formal learning – especially if they obtained a graduate degree at 30 years of age or older – but participated less frequently in informal learning (Fig 2B). Informal learning was additionally associated with a paternal graduate degree or – paradoxically – a mother who did not complete high school or GED. Replacing educational attainment before 30 and degree at 30 or older with highest educational attainment (regardless of age) did not significantly alter the model fit. Importantly, current ADI did not associate with frequency of any lifelong learning activity. Based on these findings, parental education and personality traits are more strongly associated with lifelong learning activities than current neighborhood-level SES.

### Lifelong learning influenced plasma markers related to vascular age and arthritis

While lifelong learning activities may reflect early life and intrinsic factors, their biological consequence remains unknown. We therefore next sought to determine if there existed age-related biological correlates for the four types of lifelong learning activities through aptamer-based plasma proteomics. PCA using a subset of NJHealth participants (n=226, including 164 LLS participants) revealed 34 PCs accounting for 74.1% of the variance among 100 plasma proteins at a minimum eigenvalue of 0.95 (Supplementary Datafile). Examination of these PCs revealed functionally correlated proteins representing specific inflammatory and other physiologic pathways (Supplementary Materials).

Three of the 34 PC scores showed associations with age (PC1, p<0.0001; PC10, p=0.0018; PC27, p=0.0122). PC1 scores demonstrated a parabolic rise with age (ρ=0.663, p<0.001; Fig 3A), and has top loading from multiple soluble myeloid cell surface receptors (sTREM1, sTREM2, sTNFR2, CCL23/MPIF1), related protein (sTNFR1, TIMP1), as well as a renal marker cystatin C (CST3). Regression analysis controlling for sex, race/ethnicity, medical co-morbidities, parental education, depression (PHQ8), anxiety (GAD7), ACEs, and ADI further showed positive association with traditional (diabetes, chronic kidney disease, myocardial infarction) and non-traditional (depression measured by PHQ8 score) vascular risk factors, but negative association with use of cardioprotective statins (Table 2, adjusted model R^2^=0.547). We thus considered PC1 a suitable surrogate for vascular age (VasAge). VasAge was not associated with educational attainment, parental education, ACEs, or ADI, but was negatively associated with any non-formal learning (p=0.012). There did not appear an obvious dose effect, but the small number of LLS participants with frequent non-formal learning (6 and 16 reporting daily and weekly, out of 166) and plasma proteomic data limited this interpretation. Non-formal learning’s effect was also not mediated by social gatherings (frequency of religious attendance, average number of volunteer hours) or other needs for structured learning (having a foreign accent, being a caregiver), nor diminished by restricting the analysis to those 50 years of age or older (data not shown).

**Figure 3.**
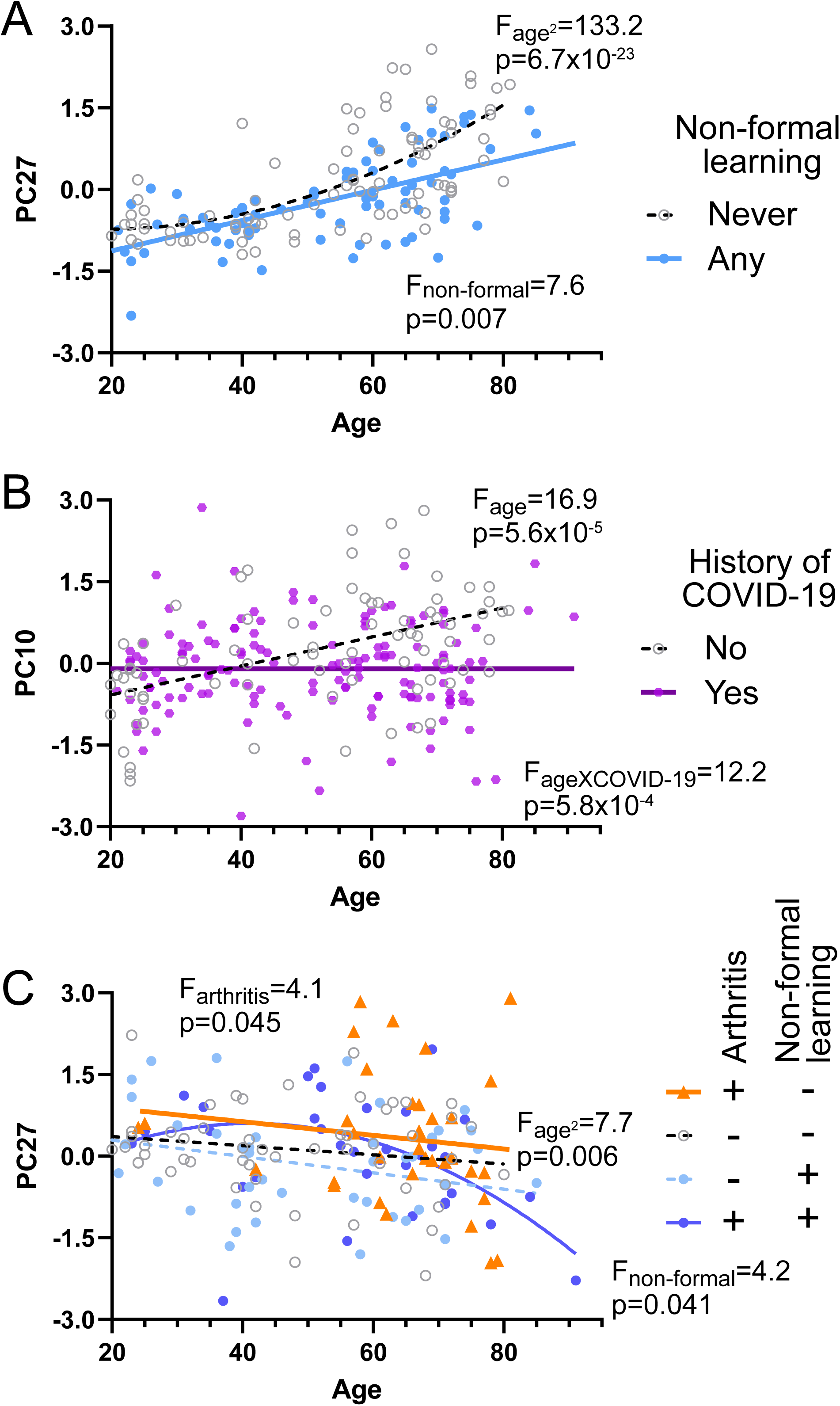
Relationship between age, plasma proteomic PCs, medical co-morbidities, and lifelong learning activities. Levels of 100 plasma proteins related to inflammation, signaling, and physiologic functions were analyzed by PCA to generate orthogonal proteomic PCs and regression for association with age. Further examinations showed associations with age in the context of vascular risks (A), COVID-19 (B), and arthritis (C). F-statistics and p-values from simple models are shown, with detailed co-variates in Tables 3 (PC1, PC27) and S9 (PC10).

**Table 2.** Lifelong learning activities’ associations with age-related PC1 and PC27. .

|  | **PC1 with early life variables**<br>**(adj R<sup>2</sup>=0.547)** |  | **PC1 with later life variables**<br>**(adj R<sup>2</sup>=0.582)** |  |
| --- | --- | --- | --- | --- |
|  | B (95% CI) | p | B (95% CI) | p |
| Age<sup>2</sup> (decades<sup>2</sup>) | 0.033 (0.027, 0.062) | <0.001 | 0.037 (0.029, 0.044) | <0.001 |
| Diabetes | 0.562 (0.272, 0.851) | <0.001 | 0.495 (0.168, 0.823) | 0.003 |
| Myocardial infarction | 0.408 (-0.068, 0.884) | 0.093 | 0.424 (-0.107, 0.955) | 0.117 |
| Chronic kidney disease | -0.790 (-1.812, 0.233) | 0.129 | -0.646 (-1.661, 0.369) | 0.210 |
| CKD X Age<sup>2</sup> | 0.042 (0.022, 0.062) | <0.001 | 0.039 (0.019, 0.059) | <0.001 |
| logPHQ8 | 0.314 (0.053, 0.575) | 0.019 | 0.402 (0.103, 0.700) | 0.009 |
| Statin | -0.373 (-0.651, -0.094) | 0.009 | -0.360 (-0.631, -0.089) | 0.009 |
| Father's highest degree |  | N.S. |  |  |
| Mother's highest degree |  | N.S. |  |  |
| Highest degree before 30 |  | N.S. |  |  |
| Highest lifetime degree |  |  |  | N.S. |
| State ADI tertile |  |  |  | N.S. |
| Any non-formal learning |  |  | -0.269 (-0.479, -0.060) | 0.012 |

|  | **PC27 with early life variables**<br>**(adj R<sup>2</sup>=0.094)** |  | **PC27 with later life variables**<br>**(adj R<sup>2</sup>=0.131)** |  |
| --- | --- | --- | --- | --- |
|  | B (95% CI) | p | B (95% CI) | p |
| Age<sup>2</sup> (decades<sup>2</sup>) | -0.015 (-0.023, -0.008) | <0.001 | -0.016 (-0.025, -0.007) | <0.001 |
| Arthritis | 0.331 (0.036, 0.627) | 0.028 | 0.386 (0.064, 0.708) | 0.019 |
| Mild TBI | -0.410 (-0.837, 0.018) | 0.060 | -0.442 (-0.917, 0.033) | 0.068 |
| Dream enactment | -1.175 (-2.320, -0.031) | 0.044 | -1.955 (-3.529, -0.380) | 0.015 |
| Dream enactment X Age<sup>2</sup> | 0.054 (0.021, 0.088) | 0.001 | 0.070 (0.029, 0.111) | 0.001 |
| Father's highest degree |  | N.S. |  |  |
| Mother's highest degree |  | N.S. |  |  |
| Highest degree before 30 |  | N.S. |  |  |
| Highest lifetime degree |  |  |  | N.S. |
| State ADI tertile |  |  |  | N.S. |
| Any non-formal learning |  |  | -0.299 (-0.596, -0.003) | 0.048 |

PC10 showed a modestly positive association with age (ρ=0.218, p=9.5 x 10^-4^), with top loading from complement 5 (C5, previously shown to increase with age in serum[52]) and C5b-C6 complex. Adjusting for covariates showed the age-associated increase in PC10 scores was only apparent in those who reported no prior history of COVID-19 (p<0.001), while those having had COVID-19 failed to demonstrate any relationship between age and PC10 scores (p=0.921, Fig 3B). PC10 scores were additionally higher in those with atrial fibrillation, and lower in those with insomnia (Table S8). PC10 was not associated with any lifelong learning activities.

Finally, PC27 had a parabolic decrease with age (ρ=-0.167, p=0.012), except for those with a self-reported history of dream enactment, and increased with history of arthritis (Table 2, adjusted model R^2^ of 0.094; Fig 3C). In keeping with the association with arthritis, it has top loading from ICAM1 and its interacting partner CD18 (but not CD11a which dimers with CD18 to form Leukocyte function associated antigen 1 [LFA1]) implicated in ischemic and synovial inflammation[53–55]. PC27 scores were also lower among participants who reported any non-formal learning (Table 2, adjusted model R^2^=0.169, Fig 3C). Similar to PC1, PC27 was not associated with educational attainment, parental education, or ADI, and non-formal learning’s effect was not mediated by social gatherings or other needs for structured learning.

## Discussion

Educational attainment can influence mid/late-life aging and disease profiles through parental SES, formal education through early adulthood, one’s own SES, or lifelong learning activities. Using a subsample within the newly founded NJHealth Study, we found great diversity in educational attainment according to timing and continuity across multiple decades beyond early life. Higher openness and less neuroticism were each associated with greater frequency of lifelong learning activities beyond formal degree/certificate programs, and early life factors such as parental education had more impact on learning outside of degree/certificate- or job-related formal learning than current ADI. Importantly, non-formal learning was associated with protective profiles against plasma proteomic markers elevated in vascular disease and arthritis, representing testable mechanisms linking early life behaviors and traits to mid-/late-life health outcomes including dementia. We discuss these findings below.

Formal education has well-acknowledged associations with cognitive aging and dementia risks from epidemiological studies and birth cohorts. However, the extended time periods necessary to ascertain linkages between early life age at peak educational attainment and late-life age at which outcomes can be reasonably assessed, coupled with dynamic evolution of formal educational systems and associated reforms, have limited these observations’ applicability to current or future populations. Many birth cohorts were also characterized by relative racial homogeneity or highly standardized educational systems,[56, 57] with significant variations in education quality (beyond years) in diverse societies[58, 59]. The neuroprotective effect from formal learning primarily came from crisscrossing temporal trends in dementia incidence and educational attainment in the U.S.[60, 61], but the same has not been reported in Japan or England despite similar educational gains[62, 63]. While our relatively new cohort will not have sufficient follow-up visits to determine cognitive trajectories for some time, we were able to identify more temporally proximate associations – between mid/late-life non-formal learning (itself influenced by educational attainment) and VasAge as well as arthritis-related age markers – by taking advantage of aptamer-based proteomic as an emerging technology. This approach has the potential of bridging the long gap from early life formal schooling to later-life cognitive aging. The inverse relationship between non-formal learning and VasAge is also at least consistent with the observation that reduced dementia incidence in the U.S. may be better accounted by declining cases of vascular dementia than AD[60]. In further support of a potential link between non-formal learning and dementia risks, the proportion of adults 25 to 64 years of age participating in job-related non-formal learning in the U.S. was steady from 1994 (38%) to 2003 (37%), rose to 55% in 2012/2015, and remained high at 51% in 2023. This contrasted with a fluctuating trajectory in England (40%, 27%, 51%, 48%) and a declining trajectory in Japan (41% in 2012/2015 to 30% in 2023)[16, 64–66], making non-formal learning a plausible mediator for the association between higher educational attainment and lower dementia incidence in the U.S. not observed in the other two countries.

The identification of up to three novel age-related plasma marker sets also opens new lines of investigation. Aptamer-based proteomics has certain advantages over traditional antibody-based detection systems as aptamers can be more reliably and artificially generated than antibodies. Aptamer-based plasma proteomics has been used to develop novel aging clocks or predictive tools related to later-life AD onset, yet the intrinsic analyte-level variability in high dimensional arrays remains a reproducibility challenge whether protein targets are captured by nucleic or amino acids. Similarly, aptamer-based capturing system cannot overcome altered antigen detection stemming from variable pre-analytical processing. Our pilot work showed pre-analytical processing to influence measurement precision for more than one third of the proteins in our panel (Fig S1B, S1C), and some – e.g., IFN-γ, C4, VEGF, CCL2/MCP1, CCL3/MIP1 – are frequent flyers in published biomarker panels. Some proteins in our new age marker sets are also susceptible to processing delays (sTREM2), one additional cycle of freeze-thawing (sTNFR1), or both (sTREM1, IL15Ra, sICAM1). However, identification of VasAge markers was supported by our rigorous pre-analytical processing control, as well as accumulating evidence implicating them in vascular disorders. Elevated plasma sTNFR1 and sTNFR2 levels have been found to correlate with plasma cholesterol levels[67], vascular dysfunction[68, 69], ischemic heart disease[70], intracerebral hemorrhage[71], and diabetic end-stage renal disease[72]. More recently, TREM1[73, 74] and TREM2 [75, 76] have both emerged as new targets of atherosclerosis, while at least one statin – pravastatin – can improve atherosclerosis via TREM1 inhibition[77]. We did not collect more detailed information on any lifelong learning subtype, and the beneficial effect of non-formal learning on VasAge could plausibly act through class-format exercises. However, exercise has been rarely found to decrease sTNFR1 and sTNFR2 levels [78]. Because plasma sTREM2 levels are also found to correlate with brain white matter hyperintensity volume[79], non-formal learning - subtype, intensity, and duration – should be further interrogated to identify dimensions most relevant to VasAge.

Non-formal learning had a similar effect on arthritis-related age markers, although the negative direction of change with advancing age makes clinical interpretation complex. On the one hand, arthritis is clearly associated with increasing plasma levels of proteins implicated in synovial inflammation. CD18 and ICAM-1 levels have been studied more frequently in rheumatoid arthritis than osteoarthritis due to more ambiguous level changes in the latter[55]. The age-associated decrease in PC27 scores observed here may thus account for the false negative finding in osteoarthritis if older participants with disease were compared to healthy volunteers of all ages. On the other hand, it is challenging to conceptualize an inflammatory process which decreases with age yet increases with age-related arthritis. One potential explanation could be immunosenescence, which again has been more frequently examined in rheumatoid arthritis than osteoarthritis[80, 81]. Immunosenescence is thought to account for some cases of sero-negative autoimmune arthritis[82, 83], and diagnosis of arthritis based on CD18 and ICAM-1 may improve the detection of late-onset autoimmune arthritis for appropriate therapies. Premature immunosenescence may similarly explain the loss of age-related increase in PC10 scores among those with COVID-19 due to the two entities’ reciprocal effects[49, 84, 85]. Therefore, introduction of these aptamer-measured proteomic biomarkers – in the setting of rigorously controlled pre-analytical processes – as intermediate outcomes (such as was done here) has the potential of enhancing the interpretation of early vs. mid/late-life effectors of aging in birth cohorts.

Distinct strengths in this study include the detailed characterization of parental, personal, and neighborhood factors surrounding formal education; rigorously processed and measured plasma proteomics widely available to the broader scientific community; and dedicated assessments of multiple learning types’ frequency in LLS. At the same time, survey-based work is not immune from biases in recruitment, recall, or social desirability. Even though over half of NJHealth participants and majority of LLS participants were derived from population-based sampling, their data were not adjusted for sample weights here. We were restricted to using ADI as the only current SES measure, while other measures such as family income relative to debts and income adjusted for education or years in the job could better characterize current SES. We also had to rely on parental education as the only metric for childhood SES. As mentioned before (and similar to the Framingham Study[60]), NJHealth did not collect information on aerobic and other forms of exercise which could potentially mediate non-formal learning’s effect on lowering PC1 scores. Similarly, no dietary information is collected even though diet can mediate the effect of non-formal learning on VasAge. Finally, only a portion of NJHealth and LLS participants had completed plasma proteomic analysis and we did not have a replication cohort (either prospectively recruited or in a pre-existing cohort with similarly obtained lifelong learning and proteomic measures), despite our technical rigor in pre-analytical processing enhancing their likelihood of replication.

In sum, we confirmed previous findings of increasing length and complexity towards peak educational attainment in a contemporary cohort, added to the literature novel age-associated markers potentially amenable to intervention through lifelong learning, and mostly early life factors associated with participation rates in adult learning outside of a degree or certificate program. We thus interpret early life formal education as a gateway – but not a direct mechanism – for reducing age-associated biological alterations, and call for greater examination of mid- and late-life factors promoting adult learning beyond immutable early life ones.

## Supporting information

Supplementary Methods, Results, Figure, and Tables

## Acknowledgements

The authors thank NJHealth Study participants for their contribution to this work. We are grateful to our scientific and community advisors for helping to shape the study to maximize its rigor and relevance for improving population health and health equity. We further acknowledge Karthik Kota, MD, Lalitha Mani, Victor Sotelo, and Neha Patel for assistance in recruitment; Mini Jomartin, DNP, for assistance in phlebotomy;

## Consent Statement

All human subjects provided signed informed consent forms in English, Spanish, Korean, or Haitian Creole according to participant preference.

## Author Roles

WTH was responsible for conception or design of the LLS, while WTH, PD, JCC were responsible for the design of NJHealth; WTH, MG, HH, SS, LLL, AN, ML, GS, SS, SB, MK, BC, PD, and JCC were responsible for acquisition, analysis, or interpretation of data; WTH, MM, and HH were responsible for drafting the initial manuscript; WTH, SS, LLL, AN, ML, GS, SS, SB, MK, BC, PD, and JCC were responsible for revising it critically for important intellectual content. All authors approved the final version and agreed to be accountable for all aspects of the work.

