## Supplementary Methods, Results, Figure, and Tables for "Structured adult learning outside school or job associates with improved plasma markers of age"

**Supplementary Results**

**Table S1.** Technical performance of SomaScan examined by adjacent aliquots, one freeze-thawing cycle, and 90 minute processing delay at room temperature.

**Table S2.** Biological variability of analytes across 3 weeks and according to fasting status.

**Table S3.** Factors associated with obtaining a degree beyond the first three decades of life.

**Table S4.** Ordinal regression analysis of higher degree/certificate-related formal learning frequency.

**Table S5.** Binary logistic regression analysis of higher job-related learning frequency.

**Table S6.** Ordinal regression analysis of higher non-formal learning frequency.

**Table S7.** Ordinal regression analysis of higher informal learning frequency.

**Table S8.** Factors associated with plasma proteomic PC10 in relationship to age

### Supplementary Methods

#### *Plasma Marker Selection and Qualification*

A custom assay of inflammatory proteins and, when known and available, their antagonists underwent technical validation at two vendors chosen from a larger pool for their willingness to disclose plating and freeze-thawing information related to their assays. This panel included 100 analytes for Vendor 1 (SomaLogic, Boulder, CO; only one thawing for analysis) and 65 corresponding analytes for Vendor 2 (up to three thawing cycles before analysis; Table S2). Samples used in this initial qualification came from a pilot experiment designed to examine six factors potentially influencing these proteins' concentrations in 15 volunteers who underwent weekly phlebotomy over four weeks: intermediate precision of assay (adjacent aliquots as technical replicates) relative to biological variability across the 15 volunteers, left vs. right arm phlebotomy, with or without overnight fasting, week-to-week variability, freeze-thawing cycles, and delays in processing (Fig S1A). Frozen plasma samples were randomized and blinded before shipping to each vendor for analysis. Vendor 1 was chosen due to greater sensitivity (0/100 and 18/65 analytes having undetectable levels in >80% of samples) and high intermediate precision (7/100 analytes having coefficients of variation [CV] > 10%; 96/100 analytes having inter-individual biological CV > intra-individual technical CV). Further analysis showed 62/100 analytes to be susceptible to additional freeze-thawing cycles (Fig S1B), 72/100 analytes to be susceptible to delays in plasma separation and processing (Fig S1C), and only 12 to be resistant to both (BPI, C1q, C5, IL-12Rb2, IL-23 p19, IL-31, LFA-1 β2/CD11, osteopontin/SPP1, TBK1, TGF-β3, TNF, sTNFR2; Table S2). Overnight fasting status did not make a significant impact on analyte levels (Fig S1D). Because two vendors analyzed the identical samples, we were also able to technically validate some analytes with detectable levels by both (Fig S1E).

**Figure S1.**

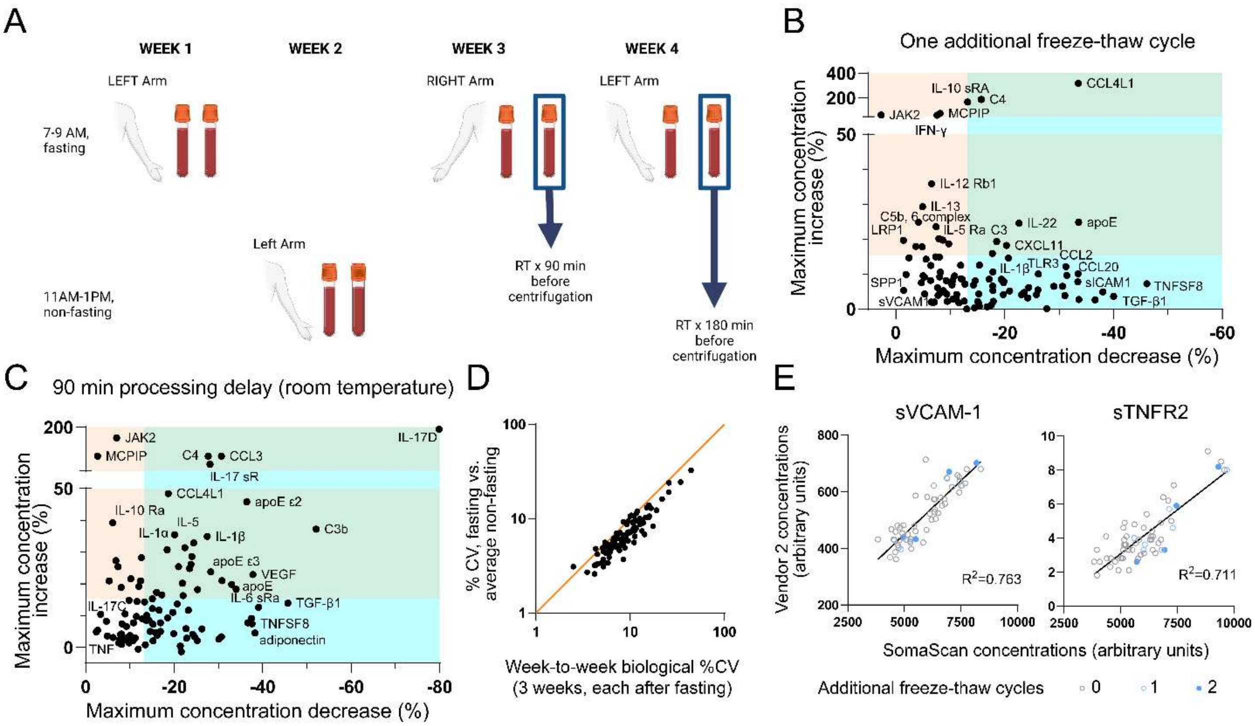


Based on these findings, ten analytes were replaced; all samples were always immediately processed (<30 minutes from arm to freezer) and shipped on dry ice without additional thawing (e.g., for sub-aliquoting).

#### *Quantitation and Statistical Analysis*

For inter-plate variability (all samples were analyzed across three 96-well plates), CVs were calculated for plate averages (in arbitrary units) of vendor-provided calibrator and quality control. Mean and standard deviation values were derived for each plasma analyte to generate level of detection (LOD), and any analyte with greater than 5% of samples below LOD were examined for a plate having outlier average levels. Only one analyte (CD6) had CV > 10% for quality control, and five (IL-7 Ra, CD6, IL-17E/IL-25, TLR3, IL-4 sR) had >5% of samples below LOD – all due to outlier blank values. Fitting each of three plates' data onto an average plate reduced the proportion of samples below LOD for IL-7 Ra (76% to 2%), CD6 (67% to 4%), IL-17E/IL-25 (19% to 0.1%), TLR3 (11% to 0), and IL-4 sR (7% to 0.4%).

Given the role of *APOE* genotype on multiple age-related disorders, we included isoforms apoE2, apoE3, and apoE4 in the panel along with total apoE. However, each isoform's levels were highly correlated with each other (Pearson's P of 0.845-0.956, p<0.001) irrespective of *APOE* genotype. Therefore, only total apoE was entered into subsequent analyses.

### Supplementary Results

***Plasma analytes form functionally correlated PCs associated with age***

Principal component analysis (PCA) of plasma analytes across 226 participants identified 34 principal components (PCs). Because majority of analytes were selected for their involvement in systemic or organ-specific inflammation, we examined membership of each PC for functional correlation using STRING-DB beyond generalized inflammation, cytokine response, or host defense. Results for the first ten PCs are shown here for illustrative purposes.

Top loading analytes for PC1 showed proteins involved in ***cardiovascular disease and acute kidney injury*** (sTNFR1, sTNFR2, TIMP1, TIMP2, NT-proBNP, cystatin C; FDR=$3.9 \times 10^{-9}$), and both PC1 and PC2 are associated with ***JAK-STAT signaling pathways*** despite only sharing IL-13 Ra1 as a top-loading protein (IL-15 Ra, gp130, IL-2, IL-13 Ra1, IL-17RC, IL-5, IL-23R for PC1, FDR=$1 \times 10^{-8}$; IL-4, IL-18, IL-4R, IL-7R, IL-13 Ra1, IL-10 Rb for PC2, FDR=$6.3 \times 10^{-10}$). PC3 has top loading from proteins involved in ***complement/C3 activation*** (C3, CRP, integrin β-2/ITGB2, FDR=$9.5 \times 10^{-4}$) and lipid storage (C3, CRP, leptin, FDR=$5.7 \times 10^{-3}$); PC4 from proteins involved in ***T cell functions*** (IL-17RD, IL-17B, IL-7, IL-10, IL-23R, IL-9, IL-4 sR, TLR4, FDR=$2 \times 10^{-10}$); PC5 from markers of T cell activation (IL-12p40, IL-23, IL22-BP, IL-9, FDR=$6.9 \times 10^{-5}$); PC6 from ***complement and coagulation cascades*** (positive loading from D-dimer and CRP, negative loading from LRP1 and kallistatin/SERPINA4, FDR=$3.3 \times 10^{-8}$); PC7 from ***leukocyte activation pathways*** (sCD5, gp130, TLR4, sTREM2, FDR=0.0124); PC8 from ***CD163 pathway*** (sCD163, plexin-B2, gp130, sVCAM-1); PC9 from ***IL-17 signaling pathway*** (negative loading from IL-6, CXCL1, IL-5, positive loading from IL-17 Rb and LRP, FDR=8.5 x $10^{-7}$); and PC10 from ***complement/C5 cascade*** (C5, C5b/C6 complex, CRP, FDR=3.4 x $10^{-4}$).

**Table S1.** Technical performance of SomaScan examined by adjacent aliquots, one freeze-thawing cycle, and 90 minute processing delay at room temperature. * Cells highlighted to reflect >10% CV between extreme value and mean value. Only 13 (out of 100) analytes were resistant to effects from freeze-thawing and processing delays.

| Protein Name | Entrez Gene Symbol | intra-run %CV, adjacent aliquots | Max reduction by freeze-thawing x 1 (%) | Max increase by freeze-thawing x 1 (%) | Range of freeze-thaw effects (as fold of %CV) | Max reduction by 90 min RT delay (%) | Max increase by 90 min RT delay (%) | Range of 90 min delay effects (as fold of %CV) |
|---|---|---|---|---|---|---|---|---|
| ACE2 | ACE2 | 2.4 | 23.6 | 4.7 | 11.81 | 9.8 | 1.4 | 4.67 |
| ADAM 17:ECD | ADAM17 | 2.8 | 4.8 | 7.5 | 4.37 | 19.9 | 4.3 | 8.64 |
| Adiponectin | ADIPOQ | 3.6 | 33.5 | 7.8 | 11.46 | 38.3 | 4.5 | 11.89 |
| apoE, pan | APOE | 8.1 | 33.6 | 24.8 | 7.22 | 34 | 18.3 | 6.46 |
| apoE ε2 | APOE ε2 | 2.0 | 8.1 | 9.2 | 8.64 | 36.4 | 45.8 | 41.10 |
| apoE ε3 | APOE ε3 | 2.3 | 5.6 | 14.3 | 8.66 | 28.3 | 23.8 | 22.65 |
| apoE ε4 | APOE ε4 | 4.9 | 36.7 | 2.6 | 8.01 | 37.5 | 9.1 | 9.51 |
| BPI | BPI | 1.6 | 7.8 | 3.9 | 7.31 | 7.5 | 1.4 | 5.56 |
| C1q | C1QA\|C1QB\|C1QC | 5.0 | 10.5 | 8.2 | 3.73 | 11.5 | 9.7 | 4.24 |
| C3 | C3 | 6.5 | 18.6 | 19.4 | 5.83 | 21.9 | 20.2 | 6.48 |
| C3b | C3 | 4.9 | 17.2 | 8.5 | 5.24 | 52.1 | 37.2 | 18.22 |
| C4 | C4A\|C4B | 12.6 | 15.7 | 187.9 | 16.16 | 27.7 | 100.4 | 10.17 |
| C5 | C5 | 4.4 | 12.2 | 8.5 | 4.71 | 8.9 | 3 | 2.70 |
| C5b, 6 Complex | C5\|C6 | 2.8 | 4.2 | 24.9 | 10.38 | 8.2 | 7.4 | 5.57 |
| CCL11/Eotaxin | CCL11 | 4.0 | 2.5 | 14.7 | 4.28 | 24.4 | 32.9 | 14.33 |
| CCL19/MIP-3β | CCL19 | 2.9 | 6.4 | 8.2 | 5.04 | 10.2 | 21.6 | 10.97 |
| CCL2/MCP-1 | CCL2 | 9.0 | 31.3 | 12.2 | 4.83 | 30.8 | 21 | 5.76 |
| CCL20/MIP-3α | CCL20 | 10.1 | 33.5 | 10.1 | 4.32 | 33 | 19.8 | 5.23 |
| CCL23/MPIF-1 | CCL23 | 2.5 | 15.3 | 4.3 | 7.82 | 9.2 | 6.6 | 6.32 |
| CCL3/MIP-1α | CCL3 | 4.0 | 17.9 | 10.6 | 7.12 | 30.7 | 100.5 | 32.80 |
| LAG-1 | CCL4L1 | 14.2 | 33.6 | 316.9 | 24.68 | 18.7 | 48.4 | 4.73 |
| sCD14 | CD14 | 6.7 | 15.4 | 12.6 | 4.18 | 12.8 | 9 | 3.25 |
| sCD163 | CD163 | 3.0 | 15.1 | 2.2 | 5.75 | 4.7 | 8.2 | 4.30 |
| CD5:ECD | CD5 | 4.2 | 23.1 | 4.1 | 6.47 | 22.4 | 31.4 | 12.81 |
| CD6 | CD6 | 4.3 | 26.2 | 5.5 | 7.36 | 11.9 | -0.6 | 2.63 |
| CXCL1/SDF-1 | CXCL1 | 5.0 | 9.8 | 18.6 | 5.68 | 15.1 | 15.3 | 6.08 |
| CXCL10/IP-10 | CXCL10 | 3.2 | 20.9 | 5.2 | 8.16 | 8.1 | 4.2 | 3.84 |
| CXCL11/I-TAC | CXCL11 | 4.5 | 20.4 | 18.2 | 8.57 | 13.8 | 10 | 5.29 |
| CXCL8/IL-8 | CXCL8 | 4.0 | 19.8 | 7.6 | 6.85 | 24.6 | 3.3 | 6.98 |
| CXCL9/MIG | CXCL9 | 1.9 | 9.5 | 2.2 | 6.18 | 15.2 | 7.3 | 11.84 |
| Progranulin/granulins | GRN | 2.4 | 6.5 | 2.0 | 3.54 | 13.7 | 8.5 | 9.25 |
| sICAM1 | ICAM1 | 5.3 | 38.1 | 5.0 | 8.12 | 30.7 | 3.3 | 6.42 |
| IFN-γ | IFNG | 9.7 | 7.6 | 62.4 | 7.22 | 9.8 | 14.8 | 2.54 |
| IL-10 | IL10 | 2.0 | 17.9 | 6.6 | 12.21 | 12.4 | 20.9 | 16.65 |
| IL-10 Ra | IL10RA | 10.6 | 13.2 | 168.2 | 17.12 | 6.1 | 39.2 | 4.27 |
| IL-10 Rb | IL10RB | 5.6 | 24.4 | 6.2 | 5.47 | 23.2 | 11.7 | 6.23 |
| IL-12 p40 | IL12B | 2.0 | 8.1 | 14.9 | 11.46 | 18.5 | 9.3 | 23.17 |
| IL-12 sRb1 | IL12RB1 | 7.1 | 6.6 | 35.9 | 5.99 | 11.6 | 14.4 | 3.66 |
| IL-12 sRb2 | IL12RB2 | 6.0 | 1.9 | 9.9 | 1.96 | 2.4 | 4.9 | 1.22 |
| IL-13 | IL13 | 4.0 | 5.0 | 29.3 | 8.57 | 24 | 28.6 | 13.15 |
| IL-13 Ra1 | IL13RA1 | 3.1 | 13.1 | 0.0 | 4.23 | 12 | 19.1 | 10.03 |
| IL-15 | IL15 | 3.2 | 6.9 | 1.8 | 2.73 | 7.3 | 25.4 | 10.22 |
| IL-15 Ra | IL15RA | 3.5 | 15.3 | 1.5 | 4.79 | 12.6 | 28.2 | 11.66 |
| IL-17 | IL17A | 3.9 | 9.4 | 4.3 | 3.51 | 21.4 | 0.4 | 5.59 |
| IL-17B | IL17B | 1.9 | 11.6 | 1.8 | 7.07 | 15.8 | 4.6 | 10.74 |
| IL-17C | IL17C | 3.6 | 8.7 | 19.8 | 7.89 | 3.4 | 10.4 | 3.83 |
| IL-17D | IL17D | 3.2 | 9.3 | 8.1 | 5.45 | 79.9 | 192.6 | 85.16 |
| IL-17F | IL17F | 3.2 | 10.4 | 7.2 | 5.50 | 22.8 | 2.9 | 8.03 |
| IL-17 sR | IL17RA | 3.3 | 14.3 | 6.9 | 6.42 | 28.1 | 72.4 | 30.45 |
| IL-17B R | IL17RB | 3.1 | 15.4 | 0.8 | 5.22 | 16.2 | 3.8 | 6.45 |
| IL-17 RC | IL17RC | 1.4 | 4.9 | 17.8 | 16.24 | 8 | 18.9 | 19.21 |
| IL-17 RD | IL17RD | 4.8 | 11.9 | 12.5 | 5.09 | 23.5 | 24.8 | 10.06 |
| IL-1α | IL1A | 4.0 | 17.9 | 5.3 | 5.80 | 18.4 | 30.7 | 12.28 |
| IL-1β | IL1B | 5.5 | 20.7 | 14.5 | 6.41 | 27.5 | 34.9 | 11.35 |
| IL-2 | IL2 | 3.5 | 10.4 | 6.9 | 4.94 | 13.3 | 14.3 | 7.89 |
| IL-21 | IL21 | 2.7 | 3.7 | 18.0 | 8.03 | 2.7 | 5.4 | 3.00 |
| IL-21R | IL21R | 1.0 | 7.1 | 2.0 | 9.10 | 26.3 | 5.8 | 32.10 |
| IL-22 | IL22 | 8.2 | 22.7 | 24.6 | 5.76 | 15.2 | 2 | 2.10 |

| | | | | | | | | |
|---|---|---|---|---|---|---|---|---|
| IL-22BP | IL22RA2 | 2.9 | 14.5 | 0.4 | 5.13 | 36.7 | 7.7 | 15.31 |
| IL-23 p19 | IL23A | 1.2 | 5.1 | 9.6 | 12.18 | 10 | 3.7 | 6.85 |
| IL-23 sR | IL23R | 3.9 | 17.6 | 1.2 | 4.81 | 16 | 17.3 | 8.54 |
| IL-17E | IL25 | 2.4 | 29.1 | 6.5 | 14.83 | 21.6 | -1.4 | 8.42 |
| IL-27 | IL27\|EBI3 | 4.4 | 27.8 | 0.1 | 6.34 | 13.5 | 0.8 | 3.25 |
| IL-2 sRa | IL2RA | 6.3 | 8.2 | 10.6 | 2.98 | 25.3 | 18.2 | 6.90 |
| IL-31 | IL31 | 3.1 | 11.2 | 4.2 | 4.97 | 10.8 | 7.7 | 5.97 |
| IL-4 | IL4 | 3.3 | 24.3 | 3.2 | 8.32 | 9.3 | 2.2 | 3.48 |
| IL-4 sR | IL4R | 3.0 | 5.3 | 9.1 | 4.80 | 14.6 | 5 | 6.53 |
| IL-5 | IL5 | 10.7 | 17.8 | 15.9 | 3.15 | 20.1 | 35.5 | 5.20 |
| IL-5 sRa | IL5RA | 5.2 | 7.4 | 23.6 | 5.97 | 6.8 | 27.3 | 6.56 |
| IL-6 | IL6 | 3.3 | 8.8 | 5.3 | 4.26 | 20.7 | 13.7 | 10.42 |
| IL-6 sRa | IL6R | 4.3 | 31.1 | 3.8 | 8.12 | 39.1 | 12.6 | 12.02 |
| gp130, soluble | IL6ST | 3.5 | 30.7 | 6.5 | 10.63 | 22.7 | 5.9 | 8.17 |
| IL-7 | IL7 | 2.3 | 12.6 | 2.3 | 6.46 | 22 | 16.3 | 16.65 |
| IL-7 sRa | IL7R | 6.3 | 25.8 | 4.2 | 4.76 | 14.3 | 8.6 | 3.63 |
| IL-9 | IL9 | 2.4 | 11.2 | 3.3 | 6.02 | 8.8 | 2.3 | 4.63 |
| LFA-1 alpha-L chain | ITGAL | 3.2 | 16.8 | 0.6 | 5.46 | 24.2 | 4.1 | 8.84 |
| LFA-1 beta-2 | ITGB2 | 2.9 | 6.5 | 12.5 | 6.53 | 6.2 | 3.1 | 3.21 |
| JAK2 | JAK2 | 15.0 | 2.7 | 63.8 | 4.07 | 7 | 163.5 | 11.37 |
| LBP | LBP | 3.4 | 7.9 | 20.2 | 8.26 | 17.2 | 16.5 | 9.91 |
| sLRP1:ECD | LRP1 | 3.2 | 1.5 | 19.7 | 6.61 | 11.2 | 2.9 | 4.41 |
| SPP1/Osteopontin | SPP1 | 2.5 | 1.5 | 5.3 | 2.72 | 8.1 | 0.9 | 3.60 |
| TBK1 | TBK1 | 3.6 | 12.0 | 8.7 | 5.75 | 8.9 | 10.5 | 5.39 |
| TGF-β1 | TGFB1 | 3.5 | 40.1 | 3.6 | 12.49 | 45.7 | 13.9 | 17.03 |
| TGF-β2 | TGFB2 | 3.7 | 18.0 | 2.2 | 5.45 | 25.2 | 2.1 | 7.38 |
| TGF-β3 | TGFB3 | 2.1 | 12.7 | 7.2 | 9.47 | 7.6 | 2.2 | 4.67 |
| TGF-β sRII | TGFBR2 | 6.2 | 33.6 | 2.7 | 5.87 | 30.1 | 2.6 | 5.27 |
| TIMP-1 | TIMP1 | 3.0 | 7.7 | 6.1 | 4.60 | 5.3 | 20.9 | 8.73 |
| TIMP-2 | TIMP2 | 6.2 | 19.9 | 8.0 | 4.49 | 19.7 | 11 | 4.95 |
| sTLR2 | TLR2 | 8.9 | 10.8 | 9.5 | 2.28 | 16.9 | 7.7 | 2.76 |
| sTLR3 | TLR3 | 4.9 | 26.2 | 10.0 | 7.40 | 16.6 | 11.7 | 5.78 |
| sTLR4 | TLR4 | 4.1 | 14.6 | 8.1 | 5.55 | 15.1 | 12.2 | 6.66 |
| TNF-α | TNF | 2.2 | 7.3 | 15.0 | 10.15 | 4.9 | 3.6 | 3.86 |
| sTNFR1 | TNFRSF1A | 3.7 | 19.4 | 8.5 | 7.55 | 10.7 | 2.1 | 3.46 |
| sTNFR2 | TNFRSF1B | 3.5 | 10.5 | 2.8 | 3.82 | 7.9 | 4 | 3.40 |
| TNFSF8 | TNFSF8 | 4.1 | 46.2 | 7.2 | 13.02 | 37.6 | 7.4 | 10.98 |
| sTREM1 | TREM1 | 5.0 | 19.8 | 4.1 | 4.78 | 21 | 25.4 | 9.28 |
| sTREM2 | TREM2 | 5.0 | 8.2 | 4.6 | 2.57 | 16.9 | 5 | 4.38 |
| sVCAM1 | VCAM1 | 2.9 | 5.4 | 4.3 | 3.32 | 23.8 | 26.1 | 17.21 |
| VEGF | VEGFA | 7.4 | 31.5 | 9.6 | 5.55 | 37.8 | 22.9 | 8.20 |
| MCPIP | ZC3H12A | 15.4 | 8.2 | 76.1 | 5.47 | 2.7 | 100.1 | 6.68 |

**Table S2.** Biological variability of analytes across 3 weeks and according to fasting status. Plasma samples generated after overnight fasting were collected weekly over three weeks, and one non-fasting sample was collected during a fourth week. Intermediate precision was calculated for the three fasting collections, and between the non-fasting and average of three fasting collections. Non-fasting did not create greater concentration variability than week-to-week variability after overnight fasting. %CV from intra-run adjacent aliquots (shaded) also included for comparison

| Protein Name | Entrez Gene Symbol | Week-to-week variablity (3 weeks, all fasting; %CV) | Fasting-NonFasting variability (%CV) | *intra-run %CV, adjacent aliquots* |
|---|---|---|---|---|
| ACE2 | ACE2 | 6.1 | 4.4 | 2.4 |
| ADAM 17:ECD | ADAM17 | 8.5 | 5.6 | 2.8 |
| Adiponectin | ADIPOQ | 15.7 | 10.3 | 3.6 |
| apoE, pan | APOE | 11.9 | 12 | 8.1 |
| apoE ε2 | APOE ε2 | 15.8 | 13.9 | 2.0 |
| apoE ε3 | APOE ε3 | 8.5 | 3.7 | 2.3 |
| apoE ε4 | APOE ε4 | 13.4 | 9.4 | 4.9 |
| BPI | BPI | 2.5 | 3.1 | 1.6 |
| C1q | C1QA\|C1QB\|C1QC | 8.8 | 4.9 | 5.0 |
| C3 | C3 | 10.7 | 9 | 6.5 |
| C3b | C3b | 14.2 | 6.8 | 4.9 |
| C4 | C4A\|C4B | 34.3 | 24.4 | 12.6 |
| C5 | C5 | 4.2 | 3.2 | 4.4 |
| C5b, 6 Complex | C5\|C6 | 5.3 | 4.5 | 2.8 |
| CCL11/Eotaxin | CCL11 | 11 | 5.6 | 4.0 |
| CCL19/MIP-3β | CCL19 | 5.4 | 5.6 | 2.9 |
| CCL2/MCP-1 | CCL2 | 10 | 10.6 | 9.0 |
| CCL20/MIP-3α | CCL20 | 11.5 | 9.5 | 10.1 |
| CCL23/MPIF-1 | CCL23 | 6 | 4.8 | 2.5 |
| CCL3/MIP-1α | CCL3 | 25.7 | 19.1 | 4.0 |
| LAG-1 | CCL4L1 | 21.8 | 16.1 | 14.2 |
| sCD14 | CD14 | 7.3 | 5 | 6.7 |
| sCD163 | CD163 | 4.3 | 3.2 | 3.0 |
| CD5:ECD | CD5 | 11.1 | 8.2 | 4.2 |
| CD6 | CD6 | 7.1 | 5 | 4.3 |
| CXCL1/SDF-1 | CXCL1 | 7.9 | 5.2 | 5.0 |
| CXCL10/IP-10 | CXCL10 | 5.2 | 3.1 | 3.2 |
| CXCL11/I-TAC | CXCL11 | 10.6 | 9.2 | 4.5 |
| CXCL8/IL-8 | CXCL8 | 9.5 | 5.8 | 4.0 |
| CXCL9/MIG | CXCL9 | 9 | 6.6 | 1.9 |
| Progranulin/granulins | GRN | 13.8 | 9 | 2.4 |
| sICAM1 | ICAM1 | 11.5 | 7.9 | 5.3 |
| IFN-γ | IFNG | 11.5 | 7.8 | 9.7 |
| IL-10 | IL10 | 10.3 | 10.8 | 2.0 |
| IL-10 Ra | IL10RA | 13.2 | 9.2 | 10.6 |
| IL-10 Rb | IL10RB | 8.4 | 6.7 | 5.6 |
| IL-12 p40 | IL12B | 9.4 | 7.1 | 2.0 |
| IL-12 sRb1 | IL12RB1 | 8.9 | 6.6 | 7.1 |
| IL-12 sRb2 | IL12RB2 | 6.3 | 4.8 | 6.0 |
| IL-13 | IL13 | 11.2 | 6.5 | 4.0 |
| IL-13 Ra1 | IL13RA1 | 7 | 6.3 | 3.1 |
| IL-15 | IL15 | 6.3 | 5.3 | 3.2 |
| IL-15 Ra | IL15RA | 11 | 10.6 | 3.5 |
| IL-17 | IL17A | 7.5 | 5.1 | 3.9 |
| IL-17B | IL17B | 6 | 3.9 | 1.9 |
| IL-17C | IL17C | 5.5 | 4.3 | 3.6 |
| IL-17D | IL17D | 44.3 | 32.5 | 3.2 |
| IL-17F | IL17F | 8.4 | 6.2 | 3.2 |
| IL-17 sR | IL17RA | 15 | 13.1 | 3.3 |
| IL-17 RB | IL17RB | 7.1 | 6.3 | 3.1 |
| IL-17 RC | IL17RC | 6.7 | 6.2 | 1.4 |
| IL-17 RD | IL17RD | 7 | 6.3 | 4.8 |
| IL-1α | IL1A | 9.8 | 7.1 | 4.0 |
| IL-1β | IL1B | 14.6 | 10.3 | 5.5 |
| IL-2 | IL2 | 6.6 | 5.8 | 3.5 |
| IL-21 | IL21 | 5.8 | 3.4 | 2.7 |
| IL-21R | IL21R | 7.7 | 4.6 | 1.0 |
| IL-22 | IL22 | 8.1 | 5.4 | 8.2 |
| IL-22BP | IL22RA2 | 11.7 | 11.4 | 2.9 |
| IL-23 p19 | IL23A | 5.6 | 4.4 | 1.2 |
| IL-23 sR | IL23R | 7.8 | 7.1 | 3.9 |
| IL-17E | IL25 | 11.1 | 7.4 | 2.4 |
| IL-27 | IL27\|EBI3 | 7 | 6.2 | 4.4 |
| IL-2 sRa | IL2RA | 12.6 | 10 | 6.3 |
| IL-31 | IL31 | 5 | 3.4 | 3.1 |
| IL-4 | IL4 | 7.4 | 5.2 | 3.3 |
| IL-4 sR | IL4R | 6.1 | 4.4 | 3.0 |
| IL-5 | IL5 | 11 | 9.3 | 10.7 |
| IL-5 sRa | IL5RA | 9.5 | 9.4 | 5.2 |
| IL-6 | IL6 | 5.7 | 5.4 | 3.3 |
| IL-6 sRa | IL6R | 13.3 | 7.4 | 4.3 |
| gp130, soluble | IL6ST | 9.6 | 7.2 | 3.5 |
| IL-7 | IL7 | 5.5 | 5.1 | 2.3 |
| IL-7 sRa | IL7R | 7.1 | 6.1 | 6.3 |
| IL-9 | IL9 | 4.5 | 4.2 | 2.4 |
| LFA-1 alpha-L chain | ITGAL | 8 | 6 | 3.2 |
| LFA-1 beta-2 | ITGB2 | 4.2 | 2.6 | 2.9 |
| JAK2 | JAK2 | 25.8 | 23.9 | 15.0 |
| LBP | LBP | 9 | 6.3 | 3.4 |
| sLRP1:ECD | LRP1 | 5.2 | 3.7 | 3.2 |
| SPP1/Osteopontin | SPP1 | 5.2 | 4.5 | 2.5 |
| TBK1 | TBK1 | 4.3 | 4.8 | 3.6 |
| TGF-β1 | TGFB1 | 19 | 12.3 | 3.5 |
| TGF-β2 | TGFB2 | 17.5 | 8.3 | 3.7 |
| TGF-β3 | TGFB3 | 4.5 | 3.3 | 2.1 |
| TGF-β sRII | TGFBR2 | 10.8 | 7.8 | 6.2 |
| TIMP-1 | TIMP1 | 7.2 | 7 | 3.0 |
| TIMP-2 | TIMP2 | 8.5 | 9.2 | 6.2 |
| sTLR2 | TLR2 | 8.4 | 5.4 | 8.9 |
| sTLR3 | TLR3 | 7 | 5.3 | 4.9 |
| sTLR4 | TLR4 | 6.5 | 5 | 4.1 |
| TNF-α | TNF | 3.5 | 2.7 | 2.2 |
| sTNFR1 | TNFRSF1A | 5.1 | 4.9 | 3.7 |
| sTNFR2 | TNFRSF1B | 7.2 | 6.5 | 3.5 |
| TNFSF8 | TNFSF8 | 16 | 10.6 | 4.1 |
| sTREM1 | TREM1 | 9.2 | 8.7 | 5.0 |
| sTREM2 | TREM2 | 8.7 | 6 | 5.0 |
| sVCAM1 | VCAM1 | 12.7 | 8.7 | 2.9 |
| VEGF | VEGFA | 10 | 7.2 | 7.4 |
| MCPIP | ZC3H12A | 16.6 | 12.9 | 15.4 |

**Table S3.** Factors associated with obtaining a degree beyond the first three decades of life. Up to bachelor's degree includes completing a bachelor's degree, associate degree, a high school (HS) diploma, or a GED.

| | **Up to bachelor's degree** | | **Graduate degree** | |
|---|---|---|---|---|
| | **O.R. (95% CI)** | **p** | **O.R. (95% CI)** | **p** |
| Female sex | 2.16 (0.87, 5.38) | 0.096 | | N.S. |
| Age (decades) | 1.39 (1.00, 1.94) | 0.052 | 1.20 (0.89, 1.62) | 0.235 |
| Education before 30 | | | | |
| HS/GED or less | Reference | | Reference | |
| Bachelor's degree | **0.033 (0.009, 0.124)** | **<0.001** | **3.68 (1.87, 7.21)** | **<0.001** |
| Master's degree | N.A. | | **4.85 (2.32, 10.15)** | **<0.001** |
| Doctorate degree | N.A. | | 1.60 (0.54, 4.73) | 0.394 |
| Big5 Agreeableness | | | | N.S. |
| 3-11 | Reference | | | |
| 12-13 | **0.27 (0.08, 0.95)** | **0.041** | | |
| 14-15 | 0.66 (0.26, 1.67) | 0.380 | | |
| Big5 Neuroticism | | N.S. | | |
| 3-5 | | | Reference | |
| 6-9 | | | **0.51 (0.31, 0.85)** | **0.010** |
| 10-15 | | | **0.55 (0.30, 0.99)** | **0.047** |
| Father having a graduate degree | | N.S. | **1.97 (1.08, 3.59)** | **0.026** |
| ACEs | | | | |
| 4 or more | Reference | | Reference | |
| 2-3 | 1.64 (0.56, 4.82) | 0.367 | 0.75 (0.05, 11.32) | 0.951 |
| 1 | **3.58 (1.07, 11.94)** | **0.038** | **17.42 (1.11, 273.0)** | **0.018** |
| None | 0.81 (0.25, 2.67) | 0.731 | 1.52 (0.12, 18.90) | 0.500 |
| ACEs X Age (decades) | | N.S. | | |
| 4 or more | | | Reference | |
| 2-3 | | | 1.01 (0.65, 1.58) | 0.806 |
| 1 | | | **0.55 (0.34, 0.90)** | **0.010** |
| None | | | 0.87 (0.58, 1.31) | 0.445 |
| Maternal education | | | | N.S. |
| <High school/GED | Reference | | | |
| High school/GED | 1.63 (0.50, 5.30) | 0.413 | | |
| Associate degree | **5.74 (1.38, 23.76)** | **0.016** | | |
| Bachelor's degree | **16.80 (2.94, 96.02)** | **0.002** | | |
| Graduate degree | **12.78 (0.95, 172.5)** | **0.055** | | |

**Table S4.** Ordinal regression analysis of higher degree/certificate-related formal learning frequency. Differences in odds are shown as proportional odds (P.O.), with p=0.180 for test of parallel line assumption and p=0.959 (> 0.05) for Pearson's goodness-of-fit test.

| | **P.O. (95% CI)** | **P** |
|---|---|---|
| **Age$^2$ (decades$^2$)** | **0.966 (0.950, 0.981)** | **<0.001** |
| **Race/ethnicity** | | |
| NHW | Reference | |
| Mexican/Chicano | 0.372 (0.045, 3.077) | 0.359 |
| Puerto Rican | 0.797 (0.104, 6.098) | 0.828 |
| Dominican | 3.873 (0.589, 25.483) | 0.159 |
| ***Other Hispanic*** | **2.609 (1.123, 6.056)** | **0.026** |
| Black/African American | 1.020 (0.369, 2.821) | 0.970 |
| Chinese | | |
| Korean | 0.376 (0.046, 3.071) | 0.361 |
| ***Asian Indian*** | **4.120 (1.216, 13.957)** | **0.023** |
| ***Other Asian*** | **5.104 (1.405, 18.541)** | **0.013** |
| More than one/Other | 0.441 (0.053, 3.651) | 0.448 |

**Table S5.** Binary logistic regression analysis of higher job-related learning frequency (weekly or daily vs less frequent). Differences in odds are shown as odds ratio (O.R.).

| | **P.O. (95% CI)** | **P** |
|---|---|---|
| **Age$^2$ (decades$^2$)** | **0.958 (0.934, 0.977)** | **<0.001** |
| **Big5 Neuroticism tertiles** | | |
| 3-5 | Reference | |
| 6-9 | 1.037 (0.551, 1.953) | 0.908 |
| **10-15** | 0.329 (0.134, 0.779) | **0.012** |
| **Big5 Openness tertiles** | | |
| 3-11 | Reference | |
| **12-13** | **3.731 (1.610, 8.621)** | **0.002** |
| **14-15** | **4.386 (1.866, 10.309)** | **<0.001** |

**Table S6.** Ordinal regression analysis of higher non-formal learning frequency. Differences in odds are shown as proportional odds (P.O.), with p=0.622 for test of parallel line assumption and p=0.268 (> 0.05) for Pearson's goodness-of-fit test.

| | **P.O. (95% CI)** | **P** |
|---|---|---|
| Female sex | 1.184 (0.772, 1.817) | 0.438 |
| Degree at 30+ | | |
| None | Reference | |
| Up to bachelor's | 0.690 (0.171, 2.783) | 0.603 |
| Graduate degree | 0.609 (0.301, 1.237) | 0.171 |
| **Female sex X Degree at 30+** | | |
| None | Reference | |
| Up to bachelor's | 3.604 (0.731, 17.796) | 0.115 |
| ***Graduate degree*** | **3.083 (1.320, 7.199)** | **0.009** |
| **Educational attainment before 30** | | |
| Doctorate | Reference | |
| Master's | 0.938 (0.464, 1.896) | 0.859 |
| ***Bachelor's*** | **0.515 (0.265, 0.997)** | **0.049** |
| Associate | 0.406 (0.093, 1.777) | 0.231 |
| ***High school/GED*** | **0.385 (0.188, 0.786)** | **0.009** |
| <High school/GED | 0.364 (0.089, 1.481) | 0.158 |
| **Big5 Openness tertiles** | | |
| 3-11 | Reference | |
| 12-13 | 1.105 (0.730, 1.675) | 0.636 |
| **14-15** | **1.179 (1.156, 2.678)** | **0.008** |
| **Insomnia** | 1.662 (1.129, 2.445) | **.010** |

**Table S7.** Ordinal regression analysis of higher informal learning frequency. Differences in odds are shown as proportional odds (P.O.), with p=0.622 for test of parallel line assumption and p=0.268 (> 0.05) for Pearson's goodness-of-fit test.

| | **P.O. (95% CI)** | **P** |
|---|---|---|
| Female sex | 0.715 (0.511, 1.001) | 0.051 |
| **Not currently married** | **2.036 (1.288, 3.219)** | **0.002** |
| **Educational attainment before 30** | | |
| Doctorate | Reference | |
| Master's | 0.792 (0.389, 1.614) | 0.521 |
| ***Bachelor's*** | 0.760 (0.386, 1.496) | 0.428 |
| Associate | **0.116 (0.028, 0.474)** | **0.003** |
| ***High school/GED*** | **0.417 (0.204, 0.854)** | **0.017** |
| <High school/GED | 0.542 (0.155, 1.891) | 0.336 |
| **Big5 Openness tertiles** | | |
| 3-11 | Reference | |
| ***12-13*** | **1.682 (1.153, 2.683)** | **0.007** |
| ***14-15*** | **2.835 (1.896, 4.238)** | **<0.001** |
| **Big5 Neuroticism tertiles** | | |
| 3-5 | Reference | |
| 6-9 | 0.921 (0.621, 1.367) | 0.686 |
| ***10-15*** | 0.511 (0.305, 0.855) | **0.011** |
| **Father with a graduate degree** | **2.296 (1.334, 3.951)** | **0.003** |
| **Maternal education** | | |
| Graduate degree | Reference | |
| Bachelor's degree | 1.484 (0.761, 2.892) | 0.246 |
| Associate degree | 0.961 (0.485, 1.906) | 0.910 |
| High school/GED | 1.600 (0.855, 2.989) | 0.141 |
| ***< High school/GED*** | **2.228 (1.089, 4.558)** | **0.028** |
| **$\log_{10}$(GAD7)** | **2.575 (1.523, 4.354)** | **<0.001** |
| Current diagnosis of asthma | 1.690 (0.965, 2.965) | 0.067 |

**Table S8.** Factors associated with plasma proteomic PC10 in relationship to age (adj $R^2$=0.158).

| Factors | B (95% CI) | p |
|---|---|---|
| Intercept | -0.770 (-1.422, -0.117) | 0.021 |
| **Age (decades)** | **0.248 (0.136, 0.360)** | **<0.001** |
| **History of COVID-19 (n=78)** | **0.897 (0.121, 1.673)** | **0.024** |
| **History of COVID X Age (decades)** | **-0.253 (-0.394, -0.113)** | **<0.001** |
| **Atrial fibrillation (n=14)** | **0.548 (0.010, 1.087)** | **0.046** |
| **Insomnia (n=52)** | **-0.343 (-0.648, -0.038)** | **0.028** |
| ACEs | | |
| Four or more (n=61) | Reference | |
| Two or three (n=47) | **-0.476 (-0.843, -0.109)** | **0.011** |
| One | -0.020 (-0.413, 0.373) | 0.921 |
| None | -0.187 (-0.523, 0.150) | 0.276 |
